# A low-cost multi-patient pressure-controlled ventilation system with individualized parameter settings

**DOI:** 10.1101/2020.04.17.20069799

**Authors:** Alcendino Cândido Jardim-Neto, Carrie E. Perlman

## Abstract

In a major health crisis, demand for mechanical ventilators may exceed supply. This scenario has led to the idea of connecting ventilation circuits in parallel to ventilate multiple patients simultaneously with the same machine. However, simple parallel connection may be harmful when the patients’ respiratory system mechanics differ. The aim of this work was to develop and test a low-cost, multi-patient, pressure-controlled ventilation system in which parameter settings could be individualized. Two types of circuits were built from polyvinyl chloride plumbing tubes and connectors, with ball valves and water columns used to control pressures. The circuits were connected to test lungs of differing compliances, ventilated in parallel at 20 cycles per minute and assessed for control error, variability and interdependency during peak inspiratory (20 to 35 cmH2O, in 5 cmH2O steps) and positive end-expiratory (5 to 20 to 5 cmH2O, in 5 cmH2O steps) pressure changes in one of the circuits. Results showed control errors lower than 1 cmH2O, a maximum standard deviation in pressure of 1.4 cmH2O and no dependency between the parallel circuits during the pressure maneuvers or a controlled disconnection/reconnection. This pressure-control system might be used to expand a commercial ventilator or, with constant gas inflow and an automated outlet valve, as a stand-alone ventilator with individually-controlled settings for multiple patients. In conclusion, the proposed solution is presented as a potentially reliable strategy for safely individualizing pressure-control parameters in a multi-patient ventilation system during a major health crisis.

## Introduction

In a major health crisis, such as a respiratory disease outbreak or an event that causes wide-spread injury, the demand for mechanical ventilators may exceed the increase in supply that can be achieved with increased production or relocation of existing ventilators. This situation may lead to fatalities among untreated patients [1,2].

To mitigate this problem, use of a single commercial ventilator for multiple patients has been suggested [3,4]. However this approach has also been criticized [5], as simple parallel connection of circuits does not allow ventilatory parameters to be individualized. When respiratory system mechanics differ between patients [6], there is a risk of administering injurious ventilation to some patients. As matching ventilation parameters to a patient’s condition improves treatment outcome and reduces mortality rate [7], individualization is essential.

Considering the possibility of limited availability of ventilators and of highly-trained medical personnel to operate them during a time of wide-spread respiratory failure [1], the aim of this work was to develop and test a simple, low-cost, multi-patient ventilation system capable of safely enabling individualization of ventilatory parameters.

## Methods

### Design and components

The design criteria were:

- The system should be constructed of low-cost off-the-shelf, widely-available components;
- The final design should be easy to build, repair and operate, at large scale, by personnel with minimal training;
- The system should enable administration of individualized peak inspiratory (PIP)/plateau pressure and positive end-expiratory pressure (PEEP) during mandatory ventilation, with a maximum error of 3 cmH2O at inspiratory and expiratory plateau pressures;
- The system should be robust, with minimal deterioration after long-term use, cleaning and storage.

The principal operating goal of regulating pressures was achieved by employing hydrostatic pressure application to open-ended tubes submerged in water columns. The system was built using ordinary polyvinyl chloride (PVC) plumbing tubes, connectors and valves as components. The ventilation circuit for each individual patient comprised two branches: an inlet branch with an airflow regulator and an open water column to control PIP and an outlet branch with a closed water column to control PEEP. The two branches joined at a Y-connection, the stem of which led to a model lung. For testing purposes, two alternative circuits were built: a simple version with only essential components, designated the “essential pressure-control circuit” and an incrementally enhanced version, designated the “preferred pressure-control circuit”.

In the essential pressure-control circuit (Figure 1), the inlet airflow regulator was a 1/2” inner diameter (ID) manually-controlled ball-valve. The PIP control tube was a 57.5 cm long x ½” diameter pipe (15.8 mm ID), with marks at 5 cm intervals, with an open lower end that was submerged below the water surface in a transparent, open reservoir containing the PIP water column. The main airflow route passed through the airflow regulator to the patient, while the PIP tube branched off at 90° from the main route. Thus airflow to the patient passed through the regulator but bypassed the PIP tube; only excess air supply passed through the PIP tube and bubbled through the PIP water column. The PEEP control tube was a 60 cm x ½” pipe, also with marks at 5 cm intervals. Its open lower end was submerged below the water surface in a transparent, closed reservoir containing the PEEP water column. A 25 cm ⨯ 1’’ (26.6 mm ID) pipe formed a port in the top of the reservoir through which the PEEP tube was inserted; an O-ring sealed the gap between the concentric tubes while enabling the PEEP tube to be raised/lowered. A nominal ¾” diameter MPTxS male adapter in series with a nominal ¾” diameter elbow connector formed the outlet from the closed PEEP reservoir. All components of the outflow branch were arranged in series such that all outflow from the circuit passed through the PEEP tube, bubbled through the PEEP water column and passed through the reservoir outlet.

**Figure 1:**
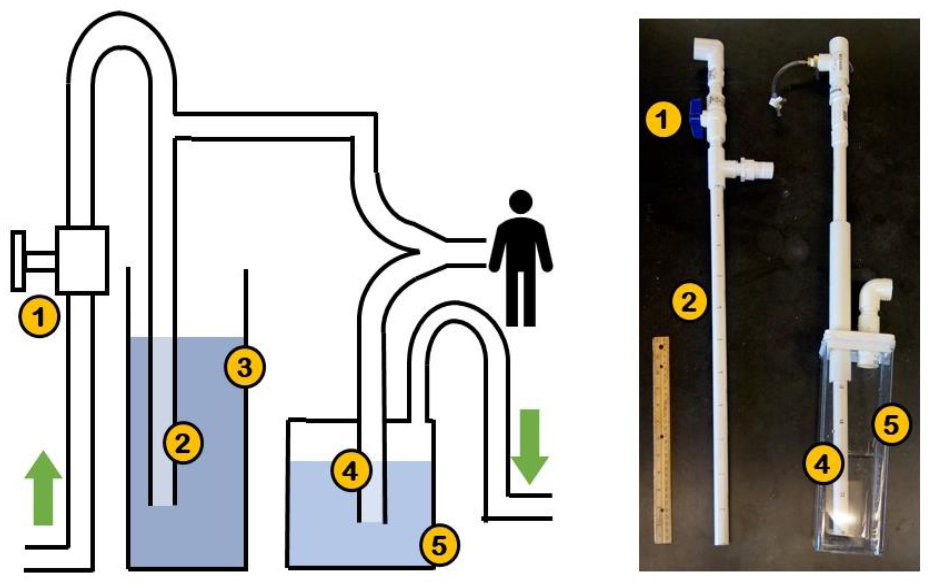
The essential pressure-control circuit. *Left and right:* Schematic and prototype, respectively. 1) Inlet airflow regulator (manually-controlled ball valve); 2) PIP control tube; 3) open PIP reservoir; 4) PEEP control tube; 5) closed PEEP reservoir. In the photograph, a 30 cm wooden ruler is shown as a reference.

The preferred pressure-control circuit (Figure 2 and Appendix) had the following additional components beyond those of the essential circuit: an inlet check valve; a PIP pressure-relief tube (60 cm ⨯ ¾” pipe) parallel to and 2.5 cm longer than the PIP control tube; a closed reservoir (6.5 ⨯ 12 ⨯ 8 cm) in series with the airflow pathway of the inlet branch and located immediately before the Y-connection to the patient, to trap any water in the inflowing air; and an outlet check valve distal to the PEEP reservoir. The inlet check valve was built from a 1-¼” coupling (42 mm ID) and a 33 mm diameter rubber ball (25.4 g), as shown in Figure 2, *right*; the outlet check valve was a commercial component (1-¼” nominal diameter inlet ⨯ 1-½” nominal diameter outlet sump pump check valve, Everbilt USA).

**Figure 2:**
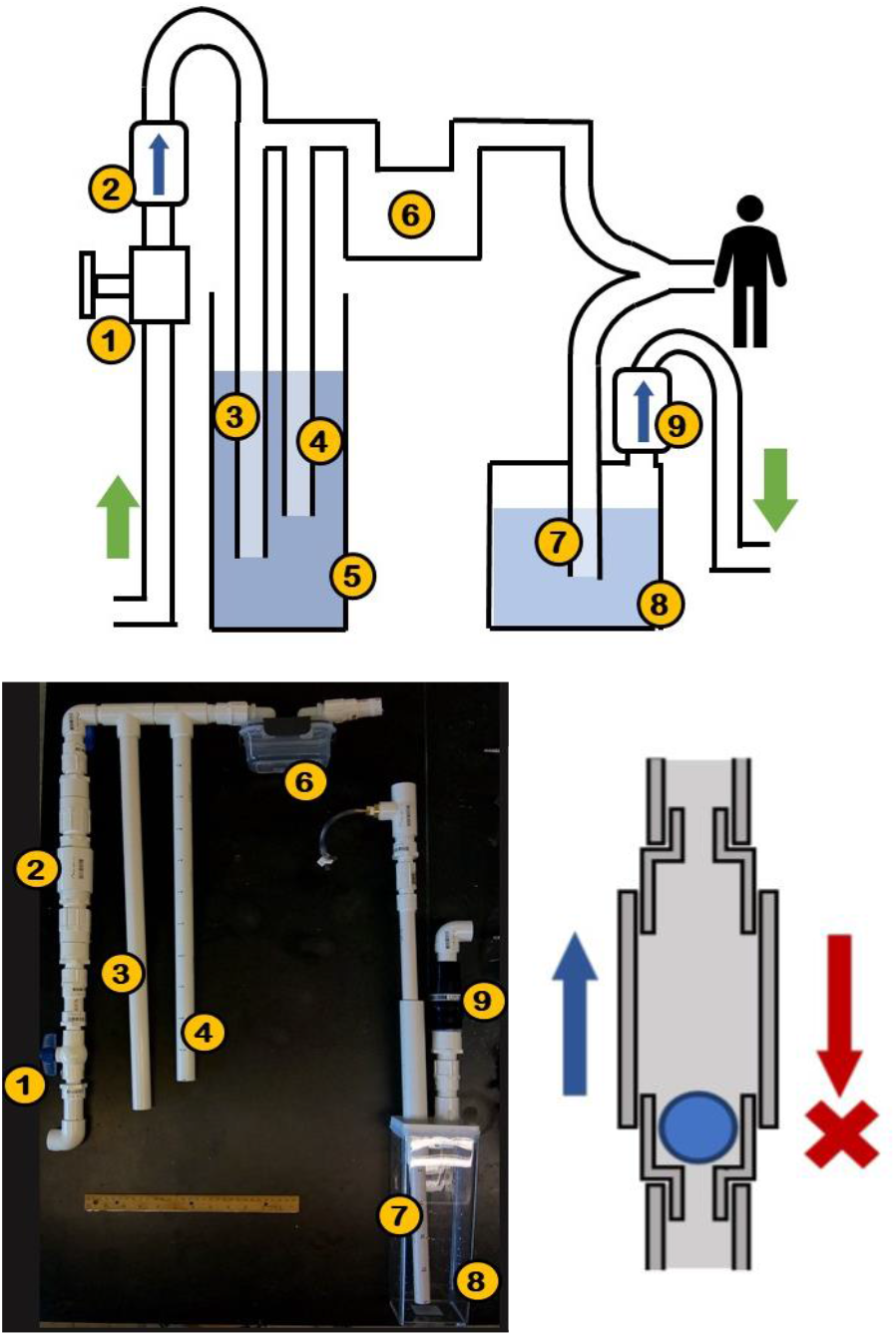
The preferred pressure-control circuit. *Top and lower left*: Schematic and prototype, respectively. 1) Inlet airflow regulator (manually-controlled ball valve); 2) inlet check valve; 3) PIP pressure-relief tube; 4) PIP control tube; 5) open PIP reservoir; 6) water trap; 7) PEEP control tube; 8) closed PEEP reservoir; 9) outlet check valve. *Lower right*: schematic of the custom-built inlet check valve (2). In the photograph, a 30 cm wooden ruler is shown as a reference

### Performance Tests

#### Setup

The test system is presented in Figure 3. The system comprised a common inlet pressure regulator (CPR) tube and then an essential circuit (circuit #1) arranged in parallel with a preferred circuit (circuit #2). The system was tested with model lungs with differing mechanics.

**Figure 3:**
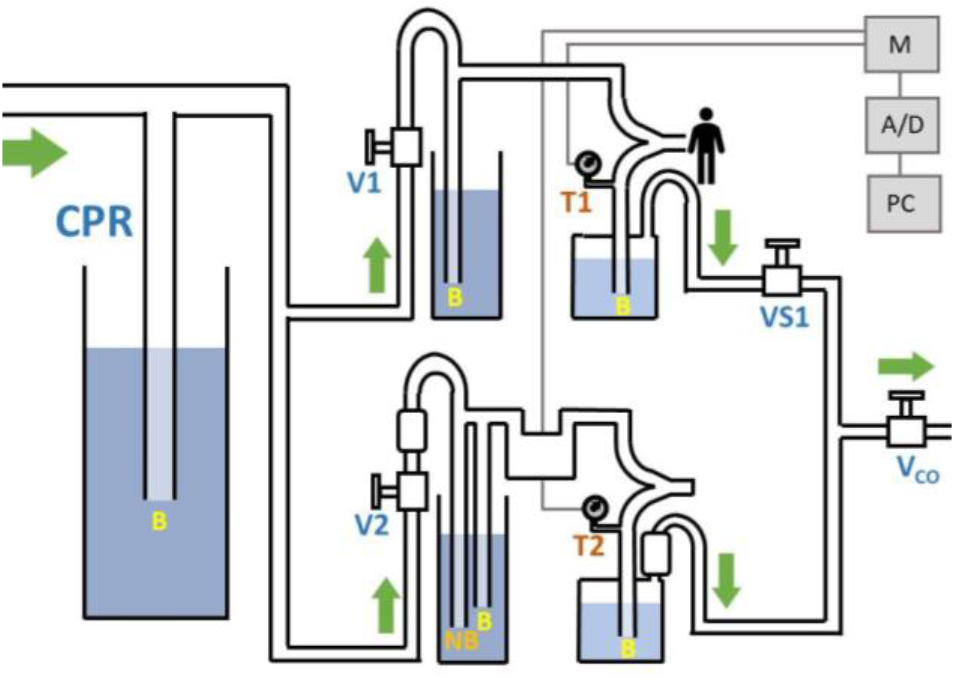
Performance test system. The air pressure from the gas source was maintained at 38 cmH2O by a common pressure regulator (CPR) tube. Then, airflow split between two parallel circuits: an “essential pressure-control circuit” (top, circuit #1), with a model test lung and an outlet safety valve (VS1), and a “preferred pressure-control circuit” (bottom, circuit #2), with a capped rigid tube used to model a low compliance respiratory system. The common outlet was passed through a ball valve (Vco) that was manually cycled on/off to achieve ventilation. As shown in previous figures, the circuits had air flow regulators (ball valves V1, V2) at their inlets. The circuits also had pressure transducers (T1, T2), located immediately before the PEEP control tubes, that connected to a signal conditioning module (M) and, in turn, a combined analog-to-digital converter/data acquisition device (A/D) and a computer (PC). Under proper operation, bubbles, “B,” exited all tubes excepting that label “NB.”

Details of the test system were as follows. Air from a gas source had its pressure regulated by the CPR. The CPR was a 60 cm x ¼” pipe that branched off at 90° from the common inspiratory line and had an open lower end submerged below the water surface in a transparent, open reservoir. Submerging the lower end by 38 cm set the common inspiratory pressure to 38 cmH2O (validated *a priori* with a calibrated pressure transducer). Circuit 1 included a model test lung (Airway Management Trainer, Laerdal, Norway), connected via a 7.0 mm endotracheal tube, and a safety valve (a ½” ID ball-valve) distal to the PEEP water column. The safety valve was normally fully open. Circuit 2 included a capped rigid tube (182 cm length x 1 cm internal diameter) as the model lung, to emulate a low-compliance respiratory system.

Ventilation was initiated with the two air flow regulators (V1 and V2 in Figure 3) closed such that all airflow passed through the CPR tube. Gradually, the airflow regulators were opened until bubbles were seen coming out of the PIP control tube in the essential circuit and the PIP control tube, but not the PIP pressure-relief tube, in the preferred circuit. Cyclic pressure variation was achieved by manually opening and closing the common outlet valve (Vco in Figure 3) at a rate of 20 cycles per minute.

#### Maneuvers

Circuit 1 was subjected to PIP change, PEEP change or controlled disconnection/reconnection from the full system. Throughout all maneuvers, the PIP of circuit 2 (PIP2) was set to 30 cmH2O and the PEEP of circuit 2 (PEEP2) was set to 16 cmH2O. All pressures of both circuits were set by submerging PIP and PEEP control tubes to depths equivalent to the desired values.

In the PIP test, the PIP of circuit 1 (PIP1) was set to 20, 25, 30 and 35 cmH2O (4 cycles each) while the PEEP of circuit 1 (PEEP1) was set to 10 cmH2O. In the PEEP test, PEEP1 was set to 5, 10, 15, 20, 15, 10, and 5 cmH2O (4 cycles each) while PIP1 was set to 35 cmH2O.

For the controlled disconnection/reconnection test, the inlet flow regulator and outlet safety valve of circuit 1 (V1 and VS1, respectively, in Figure 3), in which PIP1 was previously 35 cmH2O and PEEP1 previously 10 cmH2O, were fully closed for 4 ventilation cycles and then reopened.

### Data Acquisition

Pressure transducers (model DTXPlus, Argon Medical Devices, Singapore) were attached immediately before the PEEP control tube of each circuit. The transducers were connected to an amplifier (model ETH-256, iWorx, USA) that also applied a low-pass (5 Hz) filter to the transducer signals. The treated signals were then converted and acquired, at a sampling rate of 50 Hz, by an analog-to-digital converter/data acquisition device (USB-6008, National Instruments, USA) and displayed and recorded by a custom-developed data acquisition program written in LABVIEW (National Instruments, USA), running on a personal computer.

### Data Analysis

Data was analyzed in MATLAB R2019a (Mathworks, USA). Mean and standard deviation of pressure values were calculated for the last half of each cyclic inspiratory and expiratory plateau period. These values were considered the measured PIP and PEEP, respectively. For each 4-cycle test segment, the mean and standard deviation values from the individual cycles were averaged. Control error was calculated as the difference between set (pressure-control tube depth in water) and measured pressure values.

Inter-circuit influence during ventilatory maneuvers was calculated as Pearson’s coefficient of correlation between PIP or PEEP of the manipulated circuit and PIP or PEEP of the stable circuit, respectively.

## Results

Across all ventilation tests, control errors were −0.4 ± 0.5 and −0.2 ± 0.0 cmH2O for PIP1 and PIP 2, respectively; 0.5 ± 0.1 and 0.1 ± 0.2 cmH2O for PEEP1 and PEEP2, respectively. The maximum measured pressure signal standard deviation was 1.4 cmH2O for system 1 at end-expiration with PEEP set to 10 cmH2O.

The pressures measured in response to PIP and PEEP changes are presented in Figure 4 and summarized in Table 1. The Pearson’s coefficient of correlation revealed no significant dependency of tested variables between different circuits during PIP or PEEP change.

**Table 1:**
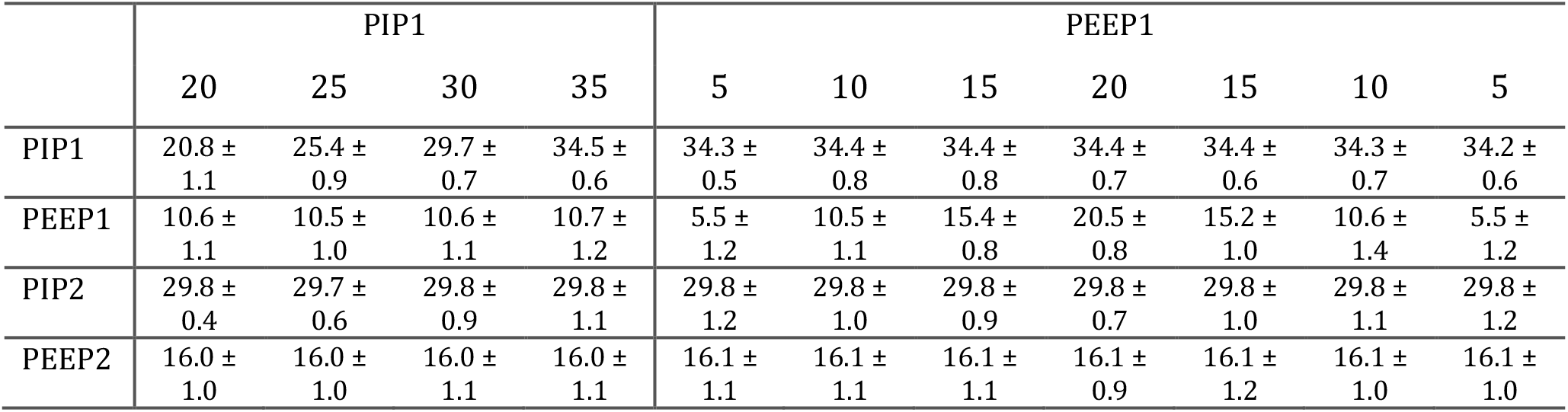
System responses to PIP and PEEP changes (mean ± standard deviation, in cmH2O)

**Figure 4:**
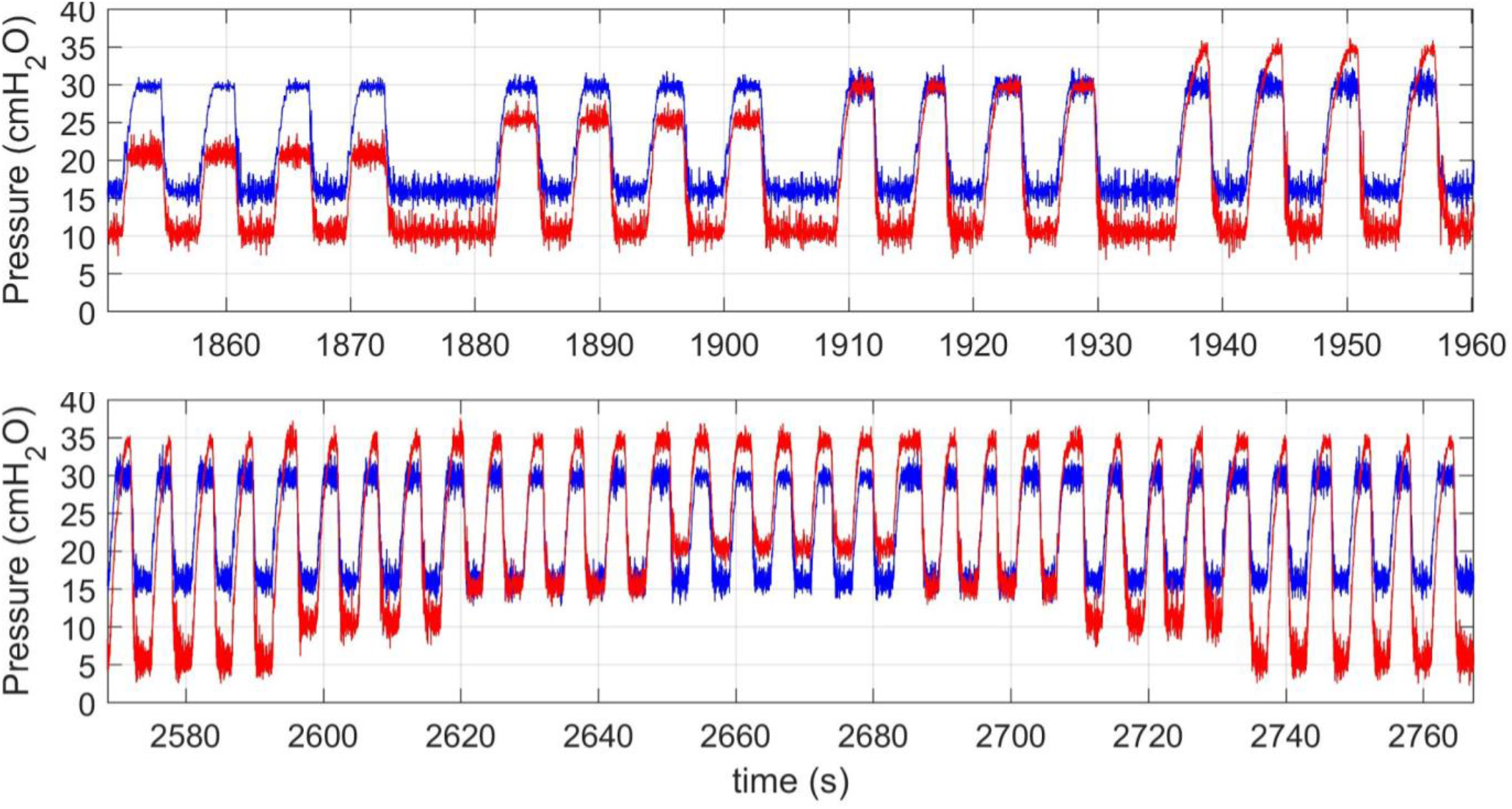
PIP/PEEP changes and inter-circuit independency. In the upper panel, PIP1 of circuit 1 (red) was set sequentially to 20, 25, 30 and 35 cmH2O, while PEEP1 was set to 10 cmH2O. In the lower panel, PEEP1 was set sequentially to 5, 10, 15, 20, 15, 10 and 5 cmH2O, while PIP1 was set to 35 cmH2O. During both tests, PIP2 and PEEP2 of circuit 2 (blue) were set to 30 and 15 cmH2O, respectively.

Controlled disconnection/reconnection of circuit 1 (Figure 5) caused no significant pressure changes in still-connected circuit 2: average PIP2/PEEP2 means and standard deviations were 29.6 ± 1.0 / 16.6 ± 1.2 cmH2O before disconnection, 29.6 ± 0.9 / 16.5 ± 1.0 cmH2O during disconnection and 29.6 ± 1.0 / 16.6 ± 1.0 cmH2O after reconnection.

**Figure 5:**
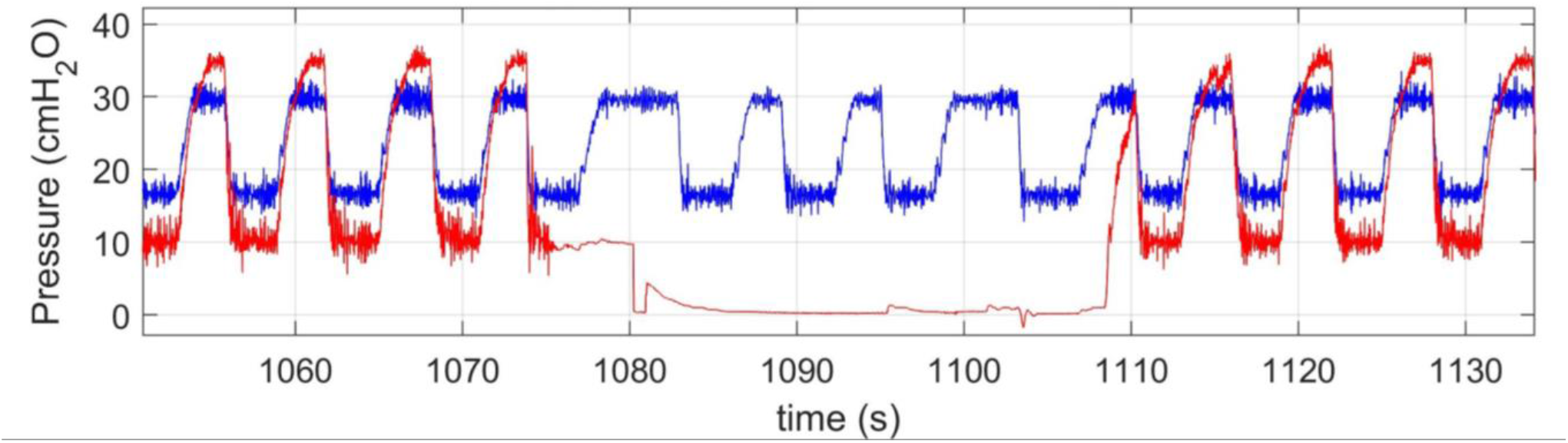
Inter-patient effects of a controlled disconnection/reconnection. Circuit 1 (red, PIP1 and PEEP1 set to 35 and 10 cmH2O, respectively) was disconnected from the overall system while parallel circuit 2 (blue, PIP2 and PEEP2 set to 30 and 16 cmH2O, respectively) was kept connected and ventilated throughout the maneuver.

## Discussion

### Working principles

The system was operated as follows. Closing Vco caused an inspiratory phase in which air bubbled out of the PIP control tubes, flowed to the model lungs and pressurized the circuits, but did not flow past the lungs or through the PEEP tubes. Opening Vco caused an expiratory phase in which the PIP control tubes stopped bubbling and air, from both the air source and the model lungs, flowed through the circuit, through the PEEP tubes – causing the PEEP tubes to bubble – and out Vco.

Airflow regulators (valves V1 and V2 in Figure 3) were required to individualize pressures at the inlets to the parallel circuits. If an ideal common pressure source (capable of providing unlimited flow) fed inflow to parallel circuits with PIP control tubes submerged to different depths, then bubbles would escape through the tubes for which hydrostatic pressures at the bottom openings were less than the common source pressure. At the top of each bubbling tube, pressure would equal the sum of the hydrostatic pressure at the bottom and the resistive pressure drop through the tube, i.e. the common source pressure. In practice, as the resistive pressure drops were negligible, it would be difficult to provide sufficient flow to raise pressure above the hydrostatic pressure associated with the least-submerged PIP tube. That is, if the source were not ideal and could provide only limited flow, then pressure would be expected to drop toward the hydrostatic pressure of the least-submerged tube and bubbles might only escape through the least-submerged tube. In either case, pressure would be the same at the entrance to each of the parallel circuits. The ball valves used as airflow regulators acted as adjustable, variable resistances that enabled air flow distribution to the different circuits to be customized. With customized resistance and airflow for each circuit, inlet pressure of each circuit was also customized. Adjusting the airflow regulator of each circuit so that bubbles just began to exit the PIP control tube of the circuit enabled the PIP control tube to maintain a steady PIP and minimized air waste. Minimizing air waste would maximize the number of patients that be ventilated in parallel.

Adjusting the airflow regulator for a circuit until bubbles were still exiting the PIP control tube at end-inspiration set the appropriate PIP for the circuit (Figures 1 and 2). That is, each PIP control tube served as both an indicator of and additional regulator for the desired PIP level. Maximum circuit pressurization was limited to the hydrostatic pressure required to expel the water column within the PIP control tube. As air pressure at the circuit inlet increased, it pushed air into and water out to the PIP control tube until the tube was completely air-filled. Air pressure in the tube was then equal to water pressure at the bottom opening of the tube. As the pressure at the base of a 1 cm water column is used as a unit of measurement (traditionally employed in respiratory physiology and clinical settings), the air pressure at that moment equaled the height of the water column from the opening at the bottom of the tube to the surface of the liquid in the reservoir, i.e. the submerged tube height. A further increase in air pressure caused air flow through the tube. Once outside the tube, the air collapsed into bubbles that traveled to the water surface such that water column height remained constant. As shown in Table 1, air pressure also remained constant at a pressure equivalent to the submerged depth of the PIP control tube, indicating that viscous loss due to flow through the tube was negligible. Thus a steady-state condition was reached, which was responsible for the inspiratory plateau phase.

The PEEP control tube operated by the same principle as the PIP control tube. However in contrast to the PIP tube, which branched off the main airflow route and acted as a release valve, the PEEP tube was positioned in-line with the main airflow route. When Vco was opened at the start of the expiratory phase, circuit pressure exceeded PEEP water column height and air flowed through the PEEP water column and out of the circuit. That measured PEEP pressures matched set PEEP pressures (Figure 4 and Table 1) demonstrated that, though the PEEP reservoir was closed, pressurized air in the top of the reservoir had a negligible effect.

### Performance evaluation

The proposed system adequately met the project’s design and assembly criteria. Only employing widely-available off-the-shelf plumbing tools and components ensured that the system could be built inexpensively and under adverse conditions by personnel with no training in biomedical or clinical engineering.

With both the essential and preferred pressure-control circuits, actual pressures reliably matched desired/set pressure values. Using tube submersion to adjust pressures resulted in a maximum pressure control error of less than 1 cmH2O, even with the intracycle pressure fluctuation cause by bubbling, and a low variation across ventilation cycles. These results strongly suggest that a system with either type of circuit could reasonably be used without pressure transducers, in a hypothetical adverse situation. Further, bubbling is a binary response that is easily identifiable: lack of bubbling in the PIP/PEEP control tubes would suggest that the patient is under-pressurized, bubbling in the PIP pressure-relief tube would suggest that the patient is over-pressurized. Both conditions could be corrected by adjusting the inlet airflow regulator; the common pressure source for all circuits would be used only to correct for major leaks in the system. If necessary, it might be possible for individuals with only minimal training to operate the system.

The high-frequency intracycle pressure variability probably resulted from cyclic air bubble formation. This variability had an acceptably low amplitude (standard deviations in Table 1). An increase in circuit volume might act as a mechanical low-pass filter and reduce the fluctuations. However, it should be considered that some degree of high-frequency oscillation could facilitate expulsion of bronchial secretions [8] and potentially, to some extent, exert the benefits of high-frequency ventilation [9, 10, 11].

As the bubbling may raise suspicion of ambient release of contaminated aerosols, it is important to mention that only clean inlet gas was released through the CPR and PIP tubes. The post-patient PEEP reservoir was closed and expired gas was only released through the common outlet valve (Vco in Figure 3), distal to which a low-resistance air filter could be placed. There was no recirculation and, although not tested, there was no reason to believe that expiratory air from one patient could contaminate another patient attached to a parallel circuit. Cross-contamination would be unlikely because: a) the common expiratory outlet would have the lowest air pressure in the system, creating a pressure gradient that drew all air flow toward it; b) the water in the PEEP reservoir would act as a barrier between a patient and the expiratory gas from all others; and c) the expiratory check valve of the preferred circuit would provide redundancy against air influx from the common expiratory line.

Another possible concern is patient water inhalation, as there is no physical barrier between the water in the PIP and PEEP control tubes and the patients’ lungs. Although this system is absolutely contraindicated for patients with significant ventilatory effort, this possibility must be considered. In the preferred circuit, there is a water trap in the inlet branch, which would catch any water escaping the PIP tube, and an expiratory check valve, which would prevent water inhalation from the PEEP tube. For the essential circuit, and as a necessary backup measure for the preferred circuit, all PIP and PEEP reservoirs should be placed below the patient bed level such that the PIP and PEEP tubes would have safety air columns above their water levels. The height difference between the water surface in a PIP or PEEP tube and the bed would be equivalent to the inhalation pressure required to raise the water column from the reservoir to the patient. Therefore, it is reasonable to recommend that water surfaces in the PIP and PEEP reservoirs be positioned at least 50 cm below the patient’s mouth.

As the presented results suggest, there were no significant functional differences between the essential and preferred pressure-control circuits. However, for the reasons previously mentioned, the preferred circuit included safety components that would be beneficial.

This performance test also presented limitations that should not be overlooked. Despite using a commercial test lung that visibly inflated and deflated with each cycle, compliance of the test lung was not known and flow and volume signals were not measured. Nonetheless, the minimal pressure differences over the ventilation cycle between the two circuits suggested that differing patient respiratory system mechanics (high compliance test lung vs. low compliance rigid tube) would be unlikely significantly to influence system performance. This point is important as, in theory, earlier outflow from the circuit of a patient with low respiratory system compliance might increase the PEEP of a patient with higher compliance on an essential circuit that lacked an expiratory check valve. In practice, however, no such effect was observed – likely due to the relatively high flow rates through both circuits. Another important limitation is that only a minimal number of PIP/PEEP pairs were tested during pressure change maneuvers. The low control error and lack of inter-patient influence during these tests suggests, but does not guarantee, that behavior would be similar under other conditions in the same pressure range.

### Clinical usage

This system is intended to be used to ventilate multiple patients in one of two possible configurations: 1) with a commercial ventilator as pressure source and cycling valve (Figure 6, *top*) or 2) with a custom gas source and stand-alone expiratory cycling valve (Figure 6, *bottom*). Although the system was not tested with the pressure controller of a commercial ventilator, manufacturer specifications indicate that ventilators have the capacity to provide sufficient pressure and inspiratory air flow [12].

**Figure 6:**
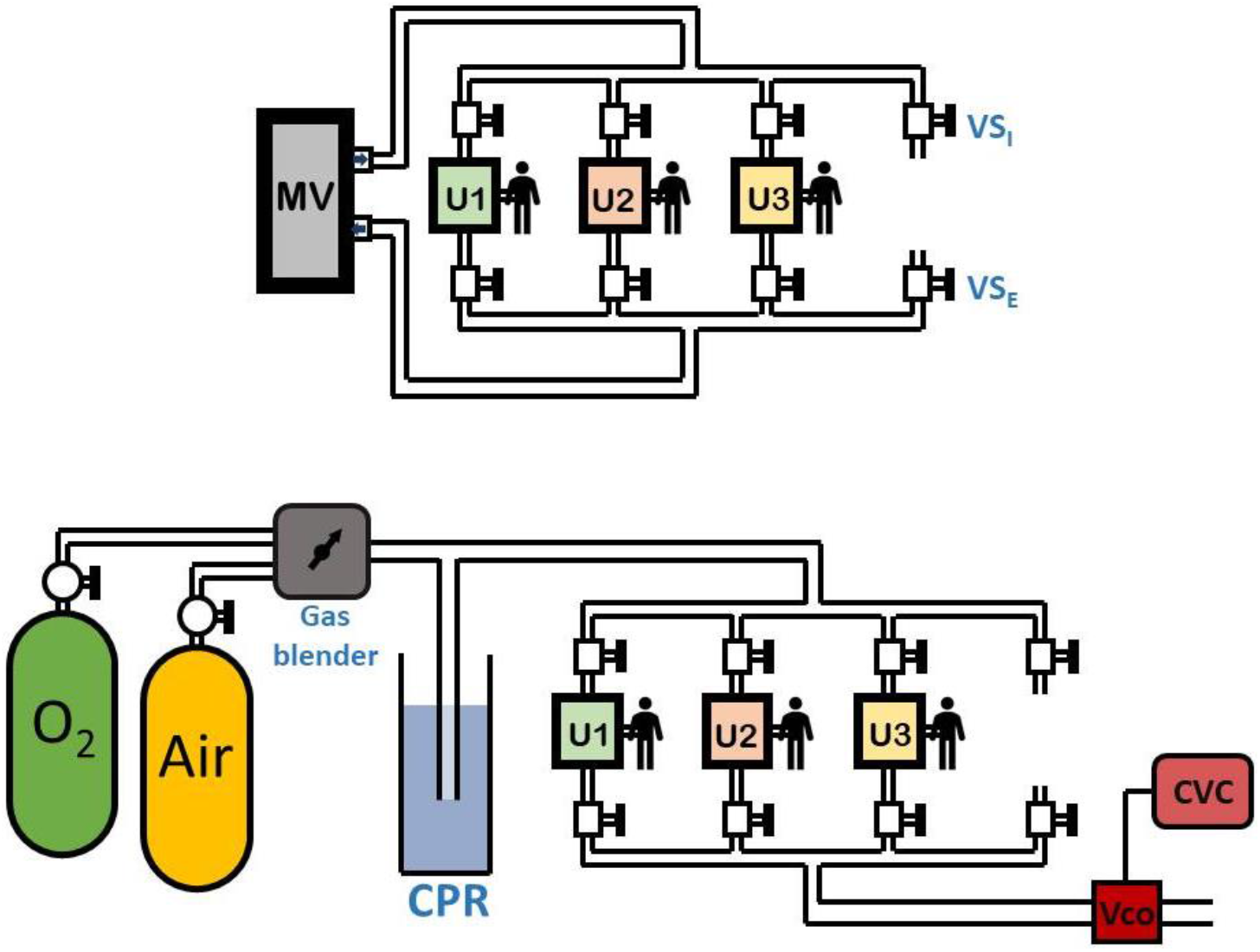
Multipatient ventilatory setups. *Top:* Commercial ventilator expansion. A single mechanical ventilator (MV) is used to ventilate three patients at the same time with individualized pressure control circuits (U1, U2 and U3). Each circuit is connected to common inspiratory and expiratory lines through inspiratory and expiratory safety valves (VSI and VSE; VSI is an additional component located proximal to the airflow regulator valves V1 and V2 in Figure 3; VSE the same as expiratory safety valve VS1 at the end of the essential circuit in Figure 3 but is an additional component at the end of the preferred circuit). The safety valves enable patients to be isolated from the common system when necessary (*far right*). *Bottom:* Multipatient custom ventilator. Oxygen and medicinal air are mixed to a desired fraction of inspired oxygen by a gas blender. Pressure is regulated by a common pressure-control tube (CPR) connected to the individual circuits (U1, U2 and U3). An automated ON/OFF valve at the common expiratory outlet (Vco) causes all patients to be ventilated at a respiratory rate determined by the valve control and power system (CVC).

Independent of the chosen configuration – expanding the capabilities of a commercial ventilator or constructing a full, custom ventilator – the recommended system requirements and operating conditions are similar:

- **<u>All</u> <u>patients</u> <u>should</u> <u>be</u> <u>sedated</u> <u>or</u> <u>paralyzed</u> <u>such</u> <u>that</u> <u>they</u> <u>lack</u> <u>significant</u> <u>ventilatory</u> <u>effort</u>** (all ventilation cycles must be mandatory);
- If using a commercial ventilator as pressure source, the ventilator must be operated in pressure-controlled ventilation mode;
- The common source pressure must be higher than the maximum required pressure among the patients;
- The common outlet must be subject to the minimum possible positive end-expiratory pressure (ideally, equal to atmospheric pressure);
- All safety valves, inspiratory and expiratory (SV_I_ and SV_E_ in Figure 6), should be fully closed on the occasion of patient disconnection (even during brief periods of time, such as for bronchial aspiration) and fully open during regular ventilation;
- All patients’ PIP control tubes must be bubbling at the end of inspiration and all PEEP control tubes at the beginning of expiration;
- Adjustments to PIP should be made during expiration and adjustments to PEEP during inspiration, as this timing makes adjustments easier;
- The water surfaces in both PIP and PEEP reservoirs must be located at least 50 cm lower than the patient mouth.

To safely connect a new patient to the system, the following steps are recommended:

1. Close both safety valves, inspiratory and expiratory;
2. Fully close the inlet airflow regulator;
3. Set the desired PIP and PEEP levels with the control tubes;
4. Open both safety valves;
5. Connect the patient’s endotracheal tube to the pressure control circuit;
6. Gradually open the inlet airflow regulator until bubbles are still seen coming out of the PIP control tube at the end of the inspiratory phase.

Although not assessed in this study, a considerable limitation of the parallel ventilation technique is the inability to individualize the patient’s respiratory rate and, therefore, CO2 washout. However, the possibility of controlling capnia by balancing the relationship between respiratory rate and ventilatory dead space should be considered. Since the respiratory rate is fixed, it is possible that all patients could be hyperventilated and expired CO2 minute-volume individually adjusted by increasing the circuit volume between the endotracheal tube and the breathing circuit Y connector.

## Conclusion

The proposed system was presented as a reliable strategy for safely individualizing pressure control parameters in a multi-patient ventilation system. With a low control error, minimal pressure fluctuations and no significant inter-patient influences, the system was also considered potentially accessible for environments with major access restrictions to complex materials or highly trained personnel.

## Data Availability

All data presented in the manuscript is available upon request.

## Acknowledgements

We thank Dr. Tam L. Nguyen for kindly providing operational support during prototype fabrication and testing. This study was supported by funding from NIH R01 HL113577.

## Appendix – Exploded assembly diagram for the preferred pressure-control circuit

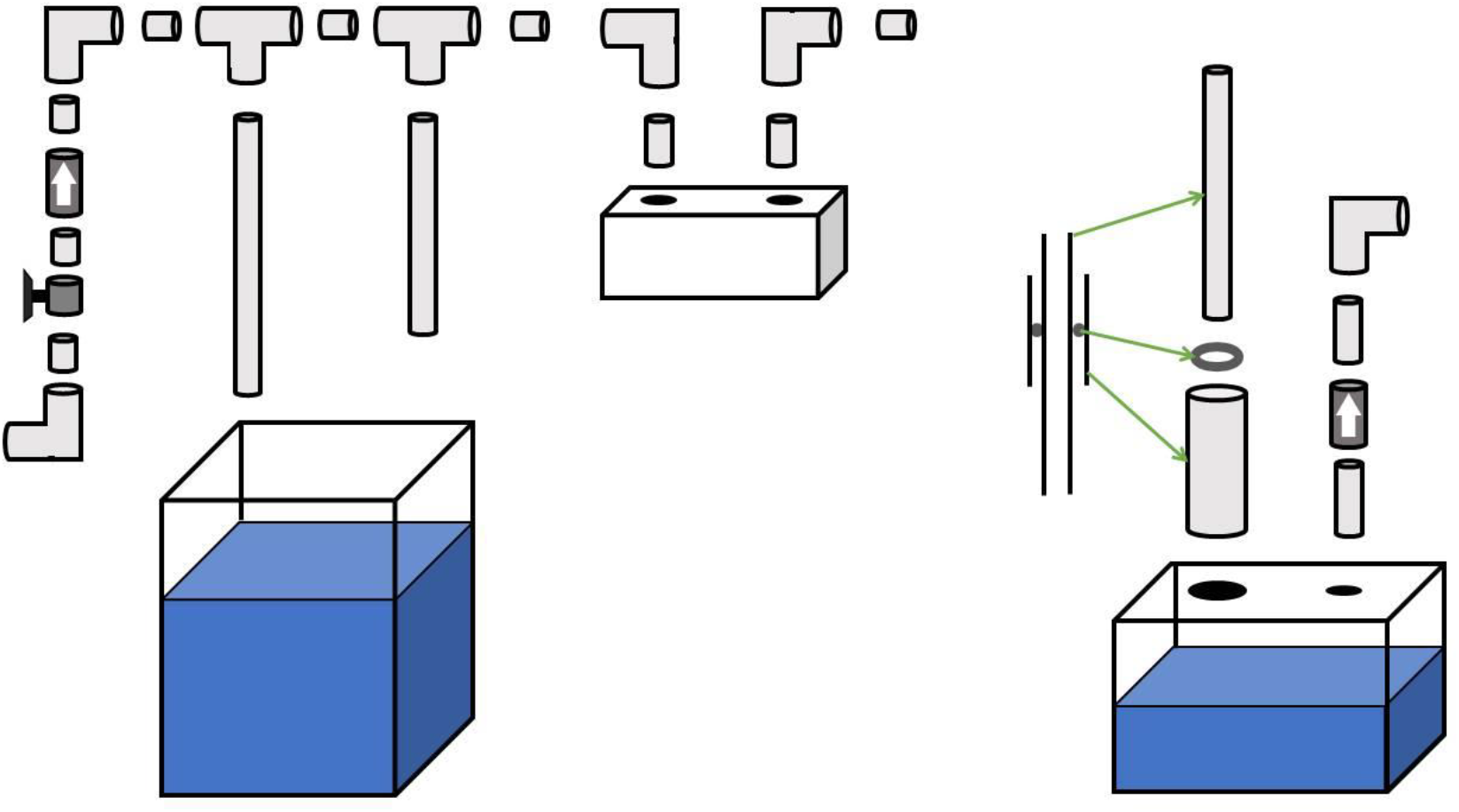

